# Estimation of the direct health costs attributable to child obesity in Brazil

**DOI:** 10.1101/2023.08.02.23293560

**Authors:** Eduardo Augusto Fernandes Nilson, Michele Gonçalves da Costa, Carolina Rocha, Olivia Honorio, Raphael Barreto

## Abstract

**Introduction:** Childhood obesity is a major global public health issue globally and in Brazil. The impacts of childhood obesity include higher risk of disease during childhood and of obesity and non-communicable diseases in adulthood and represent an important epidemiological and economic burden to countries.

**Methods:** This study is based on the modeling of total costs of hospitalizations and the additional costs attributable to childhood obesity in the Brazilian National Health System.

**Results:** The hospitalizations of children and adolescents with obesity as a primary cause totaled R$5.5 million to the Brazilian National Health System from 2013 to 2022, demonstrating that obesity is rarely considered as a cause of hospitalization especially among children and adolescents. The additional costs of hospitalizations attributable to childhood obesity totaled R$213.1 million during the same period. Considering the additional non-hospital, outpatient and medication cost attributable to childhood obesity in Brazil, the total costs were estimated at R$225.7 million in the last decade.

**Conclusion:** This study highlight that the costs of childhood obesity are not limited to the impacts on adult health and represent a relevant economic burden to the Brazilian National Health System and to families because of additional costs during childhood. Therefore, the prevention and control of childhood obesity is a public health priority that demands immediate and robust policies.

## Introduction

Childhood obesity is a major public health challenge currently. Globally, childhood obesity has increased eight times in the last four decades (1). According to the World Health Organization (WHO), in 2016, approximately 39 million children under 5 years of age were obese (2).

In Brazil, the increase of childhood obesity is also very concerning. The prevalence of obesity among children from 5 to 9 years of age increased from 2.4% in 1974 to 14.2% in 2009, while the prevalence of obesity among adolescents increased from 0.6% to 5.0% during the same period (3).

A growing of evidence has demonstrated that, compared to children with adequate weight, children with obesity have a higher risk of health issues during childhood, such as type-2 diabetes, hypertension, asthma, sleep apnea, musculoskeletal problems and metabolic disorders, as well as lower self-esteem, higher risk of bullying and lower school performance (4). Additionally, children and adolescents with obesity face higher risk of health issues in adulthood and childhood obesity is a string predictor of adult obesity and risk of non-communicable diseases such as type-2 diabetes, cardiovascular diseases, and some types of cancers. The future impacts of childhood obesity also include negative socioeconomic and workforce consequences, including reduced employability, lower productivity and salaries (5).

Considering this association between obesity during childhood and adolescence and negative outcomes in adulthood, many studies have started to project the impacts of childhood obesity on morbidity and mortality associated with comorbidities and the direct costs of treating these diseases and also the indirect costs to society related to premature mortality and retirement and losses of productivity due to absenteeism and presenteism (6)(7)(8)(9)(10).

Despite inclusive evidence on the increased risk of hospitalization among children and adolescents with obesity, strong evidence has demonstrated that childhood obesity is associated with higher risk of visits to emergency rooms (11)(12) and higher risks of hospitalization and acute infection by diseases such as influenza e Covid-19 (13)(14).

A number of studies showed that children and adolescents with obesity have higher costs of hospitalizations, outpatient procedures, prescription medications and non-hospital costs (4)(15)(16)(17)(18)(19). Based on these primary studies, a recent systematic review and meta-analysis compared the healthcare costs of children and adolescents with obesity to those with normal BMI (body-mass index) and found increased annual total medical costs attributable to childhood overweight and obesity of $237.55 per capita and annual direct and indirect costs of $13.62 billion, which were projected to reach $49.02 billion in 2050 (20)

In Brazil, there are no studies on the attributable to childhood obesity therefore the objective of this study is to estimate the direct healthcare costs of childhood obesity to the National Health System.

## Methods

This study is based on the modeling of total costs of hospitalizations and the additional costs attributable to childhood obesity in the Brazilian National Health System. The modeling is divided in the following steps: (I) estimating the percentage of additional hospitalization costs of children and adolescents with obesity compared to those with adequate BMI; (II) extracting the data on the total number and average per capita costs of hospitalizations; (III) extracting and adjustment of data on the prevalence of obesity among children and adolescents that were monitored by Primary Healthcare services; (IV) estimating the number of hospitalizations and the average per capita cost of hospitalizations among children and adolescents with obesity and those with adequate BMI; (V) estimating the total costs of hospitalizations of children and adolescents with obesity and the additional costs compared to those with adequate BMI.

Firstly, before estimating the total costs of hospitalizations of children and adolescents with obesity and the additional costs attributable to childhood obesity, we extracted the hospitalization costs which primary cause was declared as obesity in the National Hospital Information System from 2013 to 2022 by age and sex groups to analyze how childhood obesity is considered a disease by health professionals in the National Health System.

Then, as the first step for estimating the hospitalization costs attributable to childhood obesity, because there are no available studies comparing the costs of hospitalization between children and adolescents with obesity and those with adequate BMI, we estimated the percentage of additional costs among the first using a selection of the primary studies used in the meta-analysis by Ling et al, 2023, considering only the studies which assess the additional costs of hospitalizations of children and adolescents with obesity (20). We adopted this approach to avoid directly multiplying the estimated costs by the meta-analysis because of the differences between the organization and coverage of health systems and because all studies were conducted in high income countries, therefore the attributable costs could be overestimated.

Then, we extracted the average per capita costs for all hospitalizations and the total number of hospitalizations from 2013 to 2022 for each age group (1 to 4 years, 5 to 9 years, 10 to 14 years, and 15 to 19 years) from the National Hospital Information System – SIH/SUS (21).

We extracted information on the nutritional status of children and adolescents (obese and non-obese) monitored by Primary Health services using the Food and Nutrition Surveillance System (Sisvan) from 2013 to 2022, which were considered as proxies of the nutritional status of the population that predominantly uses the public health system. The data from the Sisvan system were grouped by age-group (1 to 4 years, 5 to 9 years and 10 to 19 years) and adjusted using linear regressions (22).

We conservatively assumed that children and adolescents with obesity had the same risk of hospitalization from all causes of those with adequate BMI to estimate the total number of hospitalizations for both groups according to age.

The average per capita cost of hospitalizations for children and adolescents with obesity and those with adequate BMI using the additional percentage of hospitalization costs that was previously estimated using the primary studies of the meta-analysis by Ling et al, 2023. The additional costs attributable to childhood obesity was then estimated by multiplying the additional average costs and the total number of hospitalizations of children and adolescents with obesity.

Additionally, the additional non-hospital costs, together with outpatient and medication costs, were estimated assuming that they would keep the proportionality to the additional hospitalization costs estimated by Ling et al (20).

The robustness of the model assumptions and its data inputs was assessed through sensitivity analysis changing the original model parameters and comparing the results to the primary model. The modeled scenarios included: using non-adjusted prevalences of obesity in the population, adjusting the incidence of non-communicable diseases considering the differences between Brazil and the United States of America using data from the Global Burden of Disease (GBD) Study (23) e and altering the estimated percentage of additional costs of hospitalizations in ±5%.

Further details of the parameters for the estimations and the modeling methodology are described in the Supplementary Materials.

## Results

According to information from the National Hospital Information System, show in Table 1, obesity is rarely considered as a primary cause of hospitalization of children of 1 to 9 years. For adolescents, the number of hospitalizations having obesity as a primary cause is higher, especially for those aged 15 to 19 years. For this age group, the number of hospitalizations and their corresponding costs gradually increase and peak in 2018-2019 (reaching close to R$900 thousand Reals annually), but the number of hospitalizations decreased significantly during the years of the Covid-19 pandemic (2020 and 2021) and start to increase again in 2022.

**Table 1.** Total hospitalizations and costs of hospitalization with obesity as a primary cause, Brazil 2013-2022.

| **Idade** |  | **2013** | **2014** | **2015** | **2016** | **2017** | **2018** | **2019** | **2020** | **2021** | **2022** |
| --- | --- | --- | --- | --- | --- | --- | --- | --- | --- | --- | --- |
| **1 to 4 years** | **Hospitalizations** | 0 | 1 | 1 | 0 | 0 | 1 | 0 | 1 | 0 | 1 |
| | **Cost (R\$)** | 0.00 | 61.28 | 1,161.83 | 0.00 | 0.00 | 1,156.56 | 0.00 | 55.27 | 0.00 | 486.92 |
| **5 to 9 years** | **Hospitalizations** | 0 | 0 | 1 | 2 | 3 | 1 | 0 | 3 | 3 | 1 |
| | **Cost (R\$)** | 0.00 | 0,00 | 55.27 | 6,398.84 | 488.02 | 933.85 | 0.00 | 598.49 | 11,166.65 | 390.96 |
| **10 to 14 years** | **Hospitalizations** | 4 | 2 | 2 | 3 | 2 | 2 | 4 | 2 | 2 | 4 |
| | **Cost (R\$)** | 5,668.93 | 1,251.68 | 6,446.29 | 14,311.06 | 7,013.75 | 7,088.52 | 1,804.99 | 11,365.48 | 6,051.84 | 1,723.66 |
| **15 to 19 years** | **Hospitalizations** | 86 | 72 | 100 | 128 | 143 | 143 | 147 | 38 | 24 | 43 |
| | **Cost (R\$)** | 449,984.57 | 428,119.63 | 586,050.24 | 764,832.66 | 846,466.83 | 895,592.71 | 892,080.91 | 216,468.36 | 118,523.23 | 235,168.14 |

For the estimation of the additional percentage of per capita hospitalization costs, we selected 24 of the primary studies comparing children and adolescents with obesity and those with adequate BMI primary from the meta-analysis by Ling e cols de 2023. The studies had variable sample sizes and included children and adolescents from 0 to 19 years of age, and the estimated percentage of additional hospitalization costs of children and adolescents with obesity was of 16.46% (CI 95%: 1.98%-30.94%).

The total hospitalization costs with children and adolescents with obesity increases over time, starting at R$144.7 million in 2013 and reaching R$174.0 million in 2022, and total R$1.543 billion over the last decade (CI 95% R$1.320-R$1.694 billion) (Table 2). The additional costs attributable to obesity also increase over time (R$18.9 to R$24.6 million) and total R$213.1 million (CI 95% R$186.6-R$239.6 million) from 2013 to 2022 (Table 3).

**Table 2.** Total costs of hospitalizations of children and adolescents with obesity, Brazil 2013-2020.

|  | **2013** | **2014** | **2015** | **2016** | **2017** | **2018** | **2019** | **2020** | **2021** | **2022** | **TOTAL** |
| --- | --- | --- | --- | --- | --- | --- | --- | --- | --- | --- | --- |
| **1 to 4 years** | 27,374,883 | 31,010,007 | 33,474,428 | 37,615,752 | 42,566,157 | 48,411,098 | 53,796,383 | 40,361,791 | 53,158,466 | 68,946,929 | 436,715,893 |
| **5 to 9 years** | 33,558,699 | 35,627,036 | 36,205,531 | 36,595,963 | 39,091,411 | 42,212,724 | 45,313,549 | 35,448,725 | 41,616,553 | 50,312,802 | 395,982,991 |
| **10 to 14 years** | 23,172,481 | 22,942,990 | 21,640,242 | 21,119,547 | 20,738,922 | 20,486,966 | 20,148,694 | 16,586,758 | 17,671,148 | 17,437,545 | 201,945,293 |
| **15 to 19 years** | 60,598,532 | 60,964,595 | 58,902,400 | 55,305,774 | 53,888,364 | 50,956,655 | 47,469,945 | 41,370,623 | 41,720,617 | 37,282,213 | 508,459,720 |
| **Total** | 144,704,594 | 150,544,628 | 150,222,602 | 150,637,036 | 156,284,855 | 162,067,443 | 166,728,571 | 133,767,897 | 154,166,784 | 173,979,488 | 1,543,103,897 |

**Table 3.** Estimated additional costs of hospitalizations of children and adolescents with obesity, Brazil 2013-2020.

|  | 2013 | 2014 | 2015 | 2016 | 2017 | 2018 | 2019 | 2020 | 2021 | 2022 | TOTAL |
| --- | --- | --- | --- | --- | --- | --- | --- | --- | --- | --- | --- |
| **1 to 4 years** | 3,869,315 | 4,383,123 | 4,731,458 | 5,316,815 | 6,016,532 | 6,842,688 | 7,603,874 | 5,704,955 | 7,513,707 | 9,745,334 | 61,727,801 |
| **5 to 9 years** | 4,743,369 | 5,035,719 | 5,117,487 | 5,172,673 | 5,525,393 | 5,966,576 | 6,404,864 | 5,010,516 | 5,882,310 | 7,111,485 | 55,970,391 |
| **10 to 14 years** | 3,275,325 | 3,242,887 | 3,058,750 | 2,985,152 | 2,931,352 | 2,895,739 | 2,847,926 | 2,344,463 | 2,497,736 | 2,464,718 | 28,544,047 |
| **15 to 19 years** | 8,565,326 | 8,617,068 | 8,325,586 | 7,817,219 | 7,616,875 | 7,202,491 | 6,709,660 | 5,847,549 | 5,897,019 | 5,269,671 | 71,868,464 |
| **Total** | 20,453,335 | 21,278,797 | 21,233,280 | 21,291,859 | 22,090,152 | 22,907,494 | 23,566,324 | 18,907,483 | 21,790,772 | 24,591,208 | 218,110,704 |

**Table 4.** Estimated non-hospital, outpatient and medication costs of of children and adolescents with obesity, Brazil 2013-2020.

|  | Non-hospital costs |  |  | Outpatient costs |  |  | Medication costs |  |  | Total |  |  |
| --- | --- | --- | --- | --- | --- | --- | --- | --- | --- | --- | --- | --- |
|  | Mean | CI 95% |  | Mean | CI 95% |  | Mean | CI 95% |  | Mean | CI 95% |  |
| **1 to 4 years** | 12,497,434,13 | 10,943,875,50 | 14,051,044,00 | 3,502,399,28 | 3,067,015,30 | 3,937,797,62 | 10,255,325,46 | 8,980,483,82 | 11,530,209,14 | **26,255,158,86** | 22,991,374,61 | 29,519,050,76 |
| **5 to 9 years** | 11,331,786,72 | 9,923,129,96 | 12,740,489,93 | 3,175,727,21 | 2,780,951,90 | 3,570,515,54 | 9,298,801,63 | 8,142,865,66 | 10,454,775,71 | **23,806,315,55** | 20,846,947,52 | 26,765,781,19 |
| **10 to 14 years** | 5,779,038,59 | 5,060,645,10 | 6,497,455,77 | 1,619,572,50 | 1,418,243,10 | 1,820,908,53 | 4,742,247,17 | 4,152,737,44 | 5,331,776,34 | **12,140,858,25** | 10,631,625,64 | 13,650,140,64 |
| **15 to 19 years** | 13,530,458,55 | 11,848,484,44 | 15,212,488,14 | 3,791,903,83 | 3,320,531,48 | 4,263,291,73 | 11,103,019,60 | 9,722,800,93 | 12,483,283,79 | **28,425,381,98** | 24,891,816,84 | 31,959,063,65 |
| **TOTAL** | **43,138,717,99** | **37,776,134,99** | **48,501,477,84** | **12,089,602,82** | **10,586,741,78** | **13,592,513,42** | **35,399,393,85** | **30,998,887,85** | **39,800,044,98** | **90,627,714,65** | **79,361,764,62** | **101,894,036,24** |

**Table 5.** Estimated additional non-hospital, outpatient and medication costs of children and adolescents with obesity, Brazil 2013-2020.

|  | Non-hospital costs |  |  | Outpatient costs |  |  | Medication costs |  |  | Total |  |  |
| --- | --- | --- | --- | --- | --- | --- | --- | --- | --- | --- | --- | --- |
|  | Mean | CI 95% |  | Mean | CI 95% |  | Mean | CI 95% |  | Mean | CI 95% |  |
| **1 to 4 years** | 1,766,455,36 | 1,546,866,93 | 1,986,051,04 | 445,989,35 | 390,548,32 | 501,432,21 | 1,449,543,52 | 1,269,350,46 | 1,629,742,52 | 3,661,988,23 | 3,206,765,71 | 4,117,225,76 |
| **5 to 9 years** | 1,601,696,41 | 1,402,589,20 | 1,800,810,19 | 404,391,50 | 354,121,51 | 454,663,15 | 1,314,343,23 | 1,150,956,95 | 1,477,734,90 | 3,320,431,15 | 2,907,667,66 | 3,733,208,25 |
| **10 to 14 years** | 816,840,77 | 715,299,12 | 918,385,76 | 206,233,51 | 180,596,58 | 231,871,28 | 670,295,03 | 586,970,51 | 753,622,29 | 1,693,369,30 | 1,482,866,21 | 1,903,879,33 |
| **15 to 19 years** | 1,912,468,66 | 1,674,729,28 | 2,150,215,87 | 482,854,35 | 422,830,62 | 542,880,05 | 1,569,361,22 | 1,374,273,60 | 1,764,455,27 | 3,964,684,22 | 3,471,833,50 | 4,457,551,20 |
| **TOTAL** | **6,097,461,19** | **5,339,484,53** | **6,855,462,86** | **1,539,468,71** | **1,348,097,03** | **1,730,846,69** | **5,003,543,00** | **4,381,551,53** | **5,625,554,98** | **12,640,472,90** | **11,069,133,09** | **14,211,864,53** |

During the last decade, the total non-hospital healthcare costs paid by the families, the outpatient costs and the costs of medications were estimated at a total of R$90.6 million, of which R$12.6 million represented additional costs attributable to childhood obesity. Among these costs, the healthcare costs paid by the families represent 47,6% of these additional costs (R$6.1 million).

Therefore, considering the sum of total hospital, non-hospital, outpatient, and medication costs with children and adolescents with obesity was estimated at almost R$1,6 billion from 2013 to 2022, and the additional costs attributable to childhood obesity totaled approximately R$225.7 million during the last decade.

In the sensitivity analysis, the estimated with different parameters generated small differences, between -3% and +3%, when compared to the results of the primary model.

## Discussion

The hospitalizations of children and adolescents with obesity as a primary cause totaled R$5.5 million to the Brazilian National Health System from 2013 to 2022, mostly among adolescents from 15 to 19 years. This demonstrates that obesity is rarely considered as a cause of hospitalization especially among children and adolescents.

Meanwhile, the additional costs of hospitalizations attributable to childhood obesity totaled R$213.1 million during the same period. Considering the additional non-hospital, outpatient and medication cost attributable to childhood obesity in Brazil, the total costs were estimated at R$225.7 million in the last decade. As a comparison, from 2013 to 2022, the total hospitalization costs of children and adolescents from all causes in chapter 4 of the International Code of Diseases (ICD-10), which encompasses endocrine, nutritional and metabolic diseases, including malnutrition, micronutrient deficiencies, diabetes and obesity), totaled R$156 million Reals.

These estimates demonstrate that the additional costs attributable to childhood obesity represent a large burden to the National Health System and to the families and, considering the current trends, they tend to increase in the future, together with the added risks of obesity and NCDs in adulthood, which will generate an even larger direct and indirect costs to the country. In 2018, the attributable costs of obesity among adults were estimated at R$1.42 billion (24) and future projections indicate that the direct and indirect costs of obesity may represent 6.09% of the Gross Domestic Product, totaling US$243.23 billion (25).

Additionally, if the current trends of increase in overweight and obesity in Brazil are maintained until 2030, their prevalences among adults may reach 68.1% and 29.6%, respectively (26), which will result in 5.26 million new cases and 808 thousand deaths from outcomes such as cardiovascular diseases, diabetes, cancers and chronic kidney disease until the end of the decade (27).

These trends are strongly associated with the nutritional transition in Brazil, in which traditional diets, based on foods as rice, beans, fruits and vegetables are gradually being replaced by ultra-processed products in all age groups, including children and adolescents (28). According to the National Study on Child Food and Nutrition (Enani 2019), some 80% of children under 2 years of age have already consumed ultra-processed foods, which represents a premature exposure to unhealthy foods and does not follow the recommendations of the Brazilian Dietary Guidelines for Children Under 2 Years (29). Also concerning, the results from the 2017-2018 Household Budget Survey (POF) show that the participation of ultra-processed foods in the total energy of the diets of adolescents is even higher that of the adults. i.e. 26.7% compared to 19.5% of the total calories of the diet (30).

Notably, the consumption of ultra-processed foods has been associated with higher risk of overweight and obesity (31) and of other NCDs, including cardiovascular diseases, diabetes and some types of cancers (32)(33). Also, the consumption of added sugar by Brazilians is high in all age-groups and requires effective public policy responses (30).

The results in this study are proportionally lower are lower than estimated additional costs of hospitalizations attributable to childhood obesity (20), because we assumed conservative parameters in the modeling and costs were mostly limited to the National Health System and does not include the costs of primary healthcare services. Also, if the costs of overweight were included in the estimations, the additional costs would be even higher.

The results of this study demonstrate the need for strengthening and expanding the control and prevention of childhood obesity in Brazil, considering more cost-effective interventions for treating overweight and obesity and promoting regulatory and fiscal policies that promote heathy dietary environments during all phases of childhood and adolescence, from promoting breastfeeding and complementary feeding to regulating school canteens, regulating food marketing, improving nutritional labeling, taxing ultra-processed foods and beverages and creating subsidies for fresh and minimally processed foods (34)(35)(36). In parallel, education on healthy diets should be provided for all age groups based on the recommendations of the existing national dietary guidelines (37)(38).

The strengths of this study include using a modeling method that was based on primary studies from a recent meta-analysis on the costs of childhood obesity and overweight in different countries and the use of updated data on nutritional status and costs of hospitalizations from national health information systems. The limitations of the study include the use of estimates of hospitalization costs from high-income countries for the modeling, the possible coverage and quality limitations of the national data sources, and not including primary healthcare costs and costs from the supplementary (private) health services in Brazil.

However, the assumptions that were adopted in this study allowed the first conservative estimation of direct costs of hospitalizations attributable to childhood obesity in Brazil. In the future, new studies will be important for direct and more precise assessment of these costs using data linkage of information from the national health information systems and possibly information from national childhood cohorts.

In conclusion, the results of this study highlight that the costs of childhood obesity are not limited to the impacts on adult health and represent a relevant economic burden to the Brazilian National Health System and to families because of additional costs during childhood. Therefore, the prevention and control of childhood obesity is a public health priority that demands immediate and robust policies.

## Declarations

Ethics approval and consent to participate Not applicable.

### Availability of data and materials

All relevant data on the model are publicly available. Hospitalization data is available from http://tabnet.datasus.gov.br/cgi/deftohtm.exe?sih/cnv/niuf.def and data on the nutritional status of children and adolescents is available from https://sisaps.saude.gov.br/sisvan/relatoriopublico/index

### Authors’ contributions

EAFN conceptualised the idea and was responsible for the study design and data analysis and drafted the original manuscript. All authors revised and approved the final manuscript.

## Appendix – Supplementary Materials

In the first section of the Supplementary Materials, we detail the methods for the estimation of hospitalization costs attributable to childhood obesity and, in the second section, we present tables with the parameters, input data, and intermediate results.

### Estimation of additional costs of hospitalizations attributable to childhood obesity

The percentage of additional hospitalization costs of children and adolescents with obesity was estimated using a selection of primary studies from the meta-analyses of Ling et al (20). The criteria for selecting the primary studies that were used was based on having hospitalization costs comparing children and adolescents with obesity to those with adequate Body-Mass Idex (BMI). The primary studies used for estimating the percentage of additional hospitalization costs attributable to childhood obesity and their details are listed in Table S1.

Based on these criteria, we selected 24 primary studies from six countries (United States, Canada, Australia, Germany, Netherlands and Japan), with Variable sample sizes and that, collectively analyzed children and adolescents from 0 to 19 years of age and represent local, subnational, and national analyses of hospitalization costs.

**Table S1.**
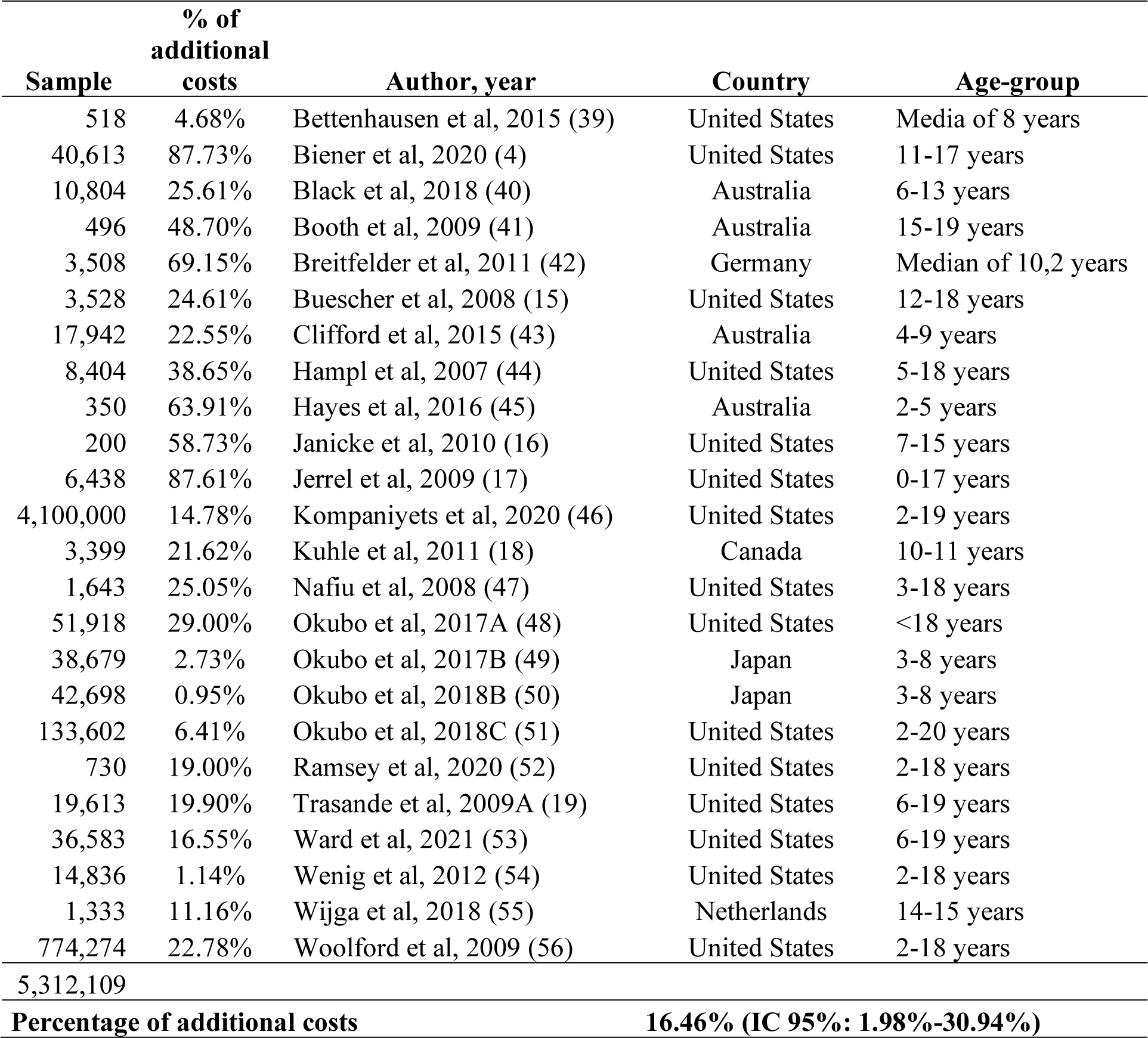
Data from the primary studies selected for the estimation of the percentage of additional hospitalization costs attributable to childhood obesity (sample size, percentage of additional costs per study, author and year of publication, country of the study, and age-group studied) and the final estimates.

| **Sample** | **% of additional costs** | **Author, year** | **Country** | **Age-group** |
| --- | --- | --- | --- | --- |
| 518 | 4.68% | Bettenhausen et al, 2015 (39) | United States | Media of 8 years |
| 40,613 | 87.73% | Biener et al, 2020 (4) | United States | 11-17 years |
| 10,804 | 25.61% | Black et al, 2018 (40) | Australia | 6-13 years |
| 496 | 48.70% | Booth et al, 2009 (41) | Australia | 15-19 years |
| 3,508 | 69.15% | Breitfelder et al, 2011 (42) | Germany | Median of 10,2 years |
| 3,528 | 24.61% | Buescher et al, 2008 (15) | United States | 12-18 years |
| 17,942 | 22.55% | Clifford et al, 2015 (43) | Australia | 4-9 years |
| 8,404 | 38.65% | Hampl et al, 2007 (44) | United States | 5-18 years |
| 350 | 63.91% | Hayes et al, 2016 (45) | Australia | 2-5 years |
| 200 | 58.73% | Janicke et al, 2010 (16) | United States | 7-15 years |
| 6,438 | 87.61% | Jerrel et al, 2009 (17) | United States | 0-17 years |
| 4,100,000 | 14.78% | Kompaniyets et al, 2020 (46) | United States | 2-19 years |
| 3,399 | 21.62% | Kuhle et al, 2011 (18) | Canada | 10-11 years |
| 1,643 | 25.05% | Nafiu et al, 2008 (47) | United States | 3-18 years |
| 51,918 | 29.00% | Okubo et al, 2017A (48) | United States | <18 years |
| 38,679 | 2.73% | Okubo et al, 2017B (49) | Japan | 3-8 years |
| 42,698 | 0.95% | Okubo et al, 2018B (50) | Japan | 3-8 years |
| 133,602 | 6.41% | Okubo et al, 2018C (51) | United States | 2-20 years |
| 730 | 19.00% | Ramsey et al, 2020 (52) | United States | 2-18 years |
| 19,613 | 19.90% | Trasande et al, 2009A (19) | United States | 6-19 years |
| 36,583 | 16.55% | Ward et al, 2021 (53) | United States | 6-19 years |
| 14,836 | 1.14% | Wenig et al, 2012 (54) | United States | 2-18 years |
| 1,333 | 11.16% | Wijga et al, 2018 (55) | Netherlands | 14-15 years |
| 774,274 | 22.78% | Woolford et al, 2009 (56) | United States | 2-18 years |
| 5,312,109 |  |  |  |  |
| **Percentage of additional costs** |  |  | **16.46% (IC 95%: 1.98%-30.94%)** |  |

### Total hospitalizations of children and adolescents

The total and average costs and the total number of hospitalizations from 2013 to 2022, according to age group (1 to 4 years, 5 to 9 years, 10 to 14 years, and 15 to 19 years), extracted from the National Hospital Information System (SIH/SUS) are detailed in Tables S2, S3 and S4.

**Table S2.**
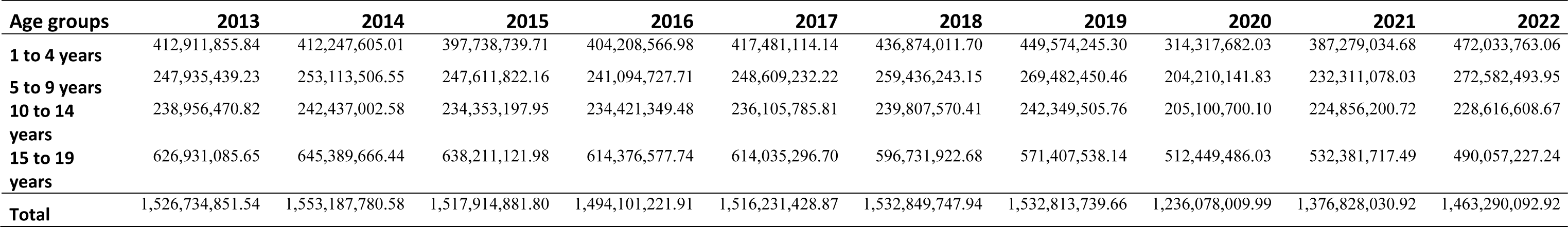
Total costs of hospitalizations from all causes of children and adolescents. by age-group. from 2013 to 2022 (National Hospital Information System - SIH/SUS).

**Table S3.**
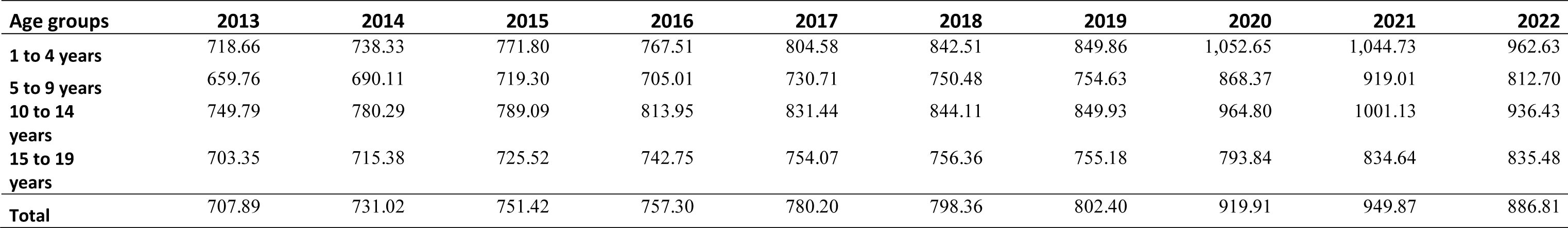
Average per capita costs of hospitalizations from all causes of children and adolescents. by age-group. from 2013 to 2022 (National Hospital Information System - SIH/SUS).

**Table S4.** Total number of hospitalizations from all causes of children and adolescents. by age-group. from 2013 to 2022 (National Hospital Information System - SIH/SUS).

| **Age groups** | **2013** | **2014** | **2015** | **2016** | **2017** | **2018** | **2019** | **2020** | **2021** | **2022** |
| --- | --- | --- | --- | --- | --- | --- | --- | --- | --- | --- |
| **1 to 4 years** | 574,300 | 557,915 | 515,036 | 526,316 | 518,652 | 518,282 | 528,792 | 298,330 | 370,538 | 490,256 |
| **5 to 9 years** | 375,595 | 366,330 | 343,774 | 341,727 | 339,939 | 345,418 | 356,797 | 234,953 | 252,722 | 335,337 |
| **10 to 14 years** | 317,838 | 309,776 | 296,165 | 287,414 | 283,594 | 283,433 | 284,571 | 212,304 | 224,430 | 243,992 |
| **15 to 19 years** | 885,986 | 897,765 | 876,668 | 824,681 | 812,366 | 786,626 | 754,426 | 643,534 | 635,544 | 584,664 |
| **Total** | 2,153,719 | 2,131,786 | 2,031,643 | 1,980,138 | 1,954,551 | 1,933,759 | 1,924,586 | 1,389,121 | 1,483,234 | 1,654,249 |

### Prevalence of obesity among Brazilian children and adolescents

The prevalence of obesity in children and adolescents was obtained from the National Food and Nutrition Surveillance System (Sisvan) from 2013 to 2022 and the adjusted prevalence through linear regression,

**Table S5.**
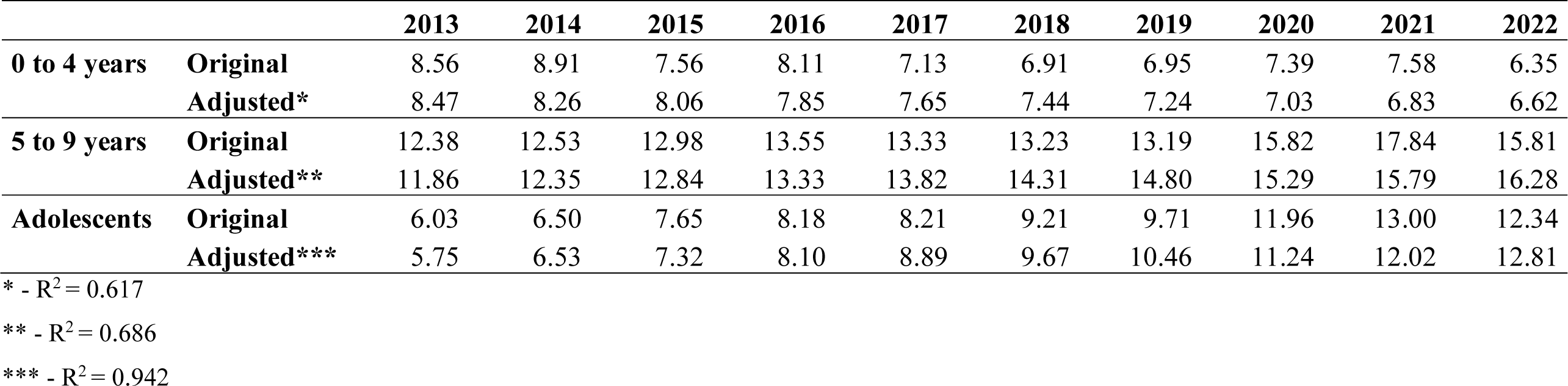
Original and adjusted prevalence of obesity by age-group from 2013 to 2022 (Sisvan).

### Estimation of the number of hospitalizations according to BMI by age-group

We used the prevalence of children and adolescents with obesity and those with adequate BMI and conservatively assumed that both groups have similar risk of hospitalization from all-causes.

We used the following formula for the estimation:

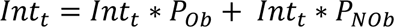

Onde:

Int_t_ = total number of hospitalizations

P_Ob_ = prevalence of children and adolescents with obesity

P_NOb_ = prevalence of children and adolescents without obesity

### Estimation of the average per capita costs of hospitalizations by nutritional status

Firstly, we estimated the average per capita cost of hospitalizations for each group (children and adolescents with and without obesity), considering the estimated percentage of additional hospitalization costs (Table S1).

Finally, the average costs of hospitalizations were multiplied by the total number of hospitalizations for the total costs of hospitalizations and the difference between the average per capita costs of hospitalizations of children and adolescents with and without obesity was used to calculate the additional costs attributable to childhood obesity.

### Proportion of non-hospital, outpatient and medication costs

The results of the meta-anlaysis by Ling et al (20), was used to estimate the proportion of non-hospital, outpatient and medication costs in relation to the hospitalization costs and used these proportions to multiply the total and additional costs (Table S6), assuming that the same proportionality would be applied to the Brazailian population.

**Table S6.** Proportion of non-hospital, outpatient and medication costs in relation to the hospitalization costs from Ling et al, 2023.

| Non-hospital costs | Outpatient costs | Medication costs |
| --- | --- | --- |
| 2.86% | 0.80% | 2.35% |

### Sensibility analysis

We performed a sensitivity analysis to assess the effect of the model assumptions and the primary inputs, by changing the input parameters of the model and comparing them to the estimates of the primary model (Figure 1). The scenarios include using non-adjusted prevalences of obesity in the population, adjusting the incidence of non-communicable diseases considering the differences between Brazil and the United States of America using data from the Global Burden of Disease (GBD) Study (23) e and altering the estimated percentage of additional costs of hospitalizations in ±5%.

**Figure 1.**
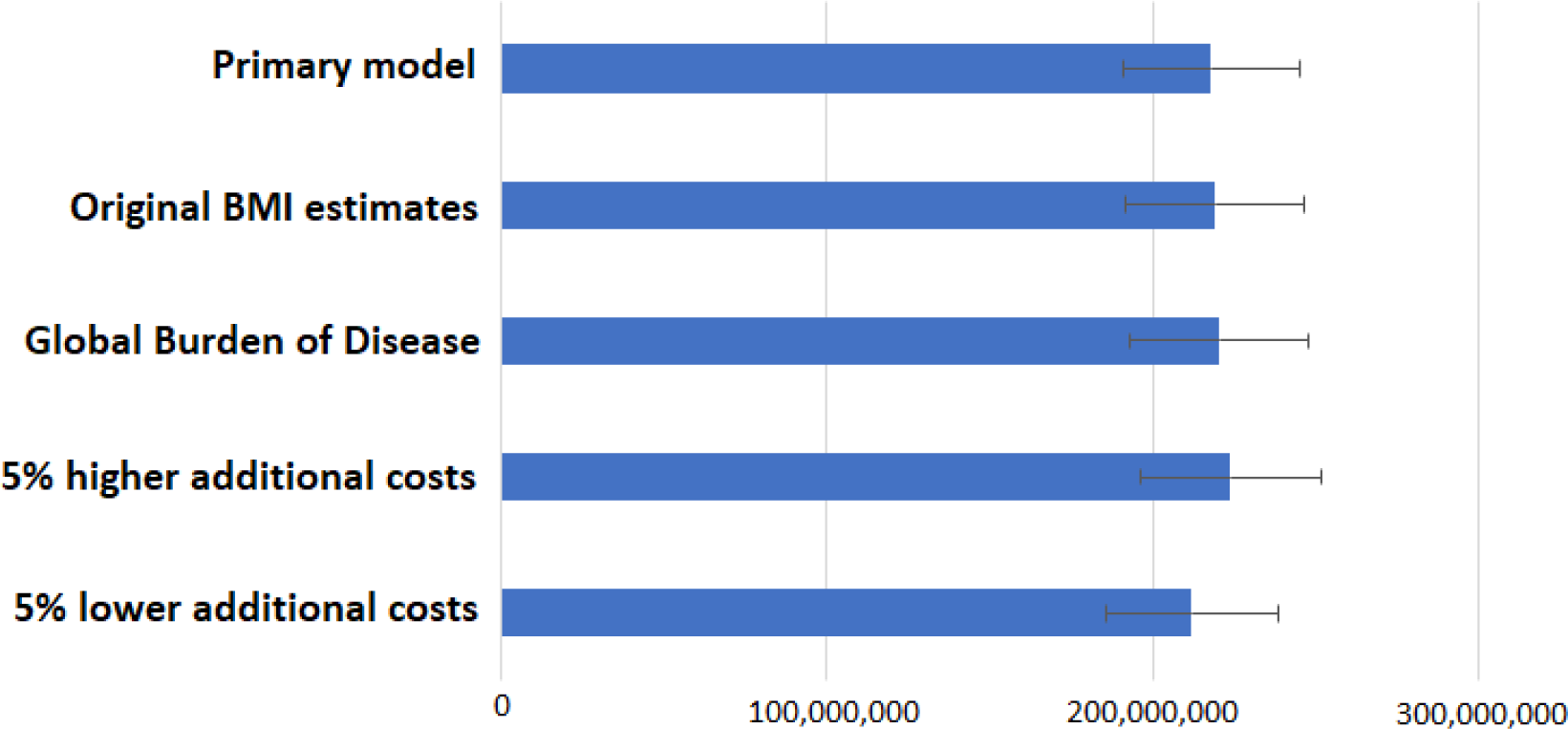
Sensitivity analysis comparing the primary model to different data input scenarios.

## Data Availability

All data produced in the present study are available upon reasonable request to the author

http://tabnet.datasus.gov.br/cgi/deftohtm.exe?sih/cnv/niuf.def

